# Improving the quality of anthropometric measures during medical consultations with children aged under five years old in Burkina Faso

**DOI:** 10.1101/2021.05.25.21257453

**Authors:** Aziza Merzouki, Wessel Valkenburg, Marc Bayala, Maroussia Roelens, Olivia Keiser, Amara Amara

## Abstract

**Objective:** Millions of medical consultations are conducted each year in Burkina Faso using the Electronic Register of Consultations (REC). Based on the consultation data collected, we present a method to quantify the quality of individual and ensembles of consultations conducted by frontline healthcare workers (FHWs).

**Methods:** We focus on anthropometric measurements and vital signs (age, weight, height, mid-upper arm circumference and temperature) of children aged between two months and five years old. We compare individual and ensemble of consultations to a multivariate probability distribution defined by an external population-specific, gold standard consultation dataset. By comparing the distributions of consultations to the reference probability distribution, we define a score to rate the quality of measurements and data entry of each FHW.

**Findings:** The defined scores allow us to detect which measurements are most problematic. They also allow us to detect potential biases in the consultation and treatment of different patient groups. No systematic gender-bias was found among FHWs. Height measurements were the most challenging; consultations with the lowest scores were associated with underestimated heights in children. Among these consultations, height was found to be even more underestimated among boys than girls.

**Conclusion:** Our findings enable us to support capacity building of frontline healthcare workers. The REC can be enriched with real-time specific alert on errors, individual FHW can be proposed targeted trainings, and dynamic dashboards can support district managers to navigate the entire population of FHWs and understand which problems should be prioritised.

**Research in context:** *Knowledge before this study:* The use of the Electronic Register of Consultations (REC) improved Frontline Healthcare Workers’ (FHWs) adherence to the Integrated Management of Childhood Illness (IMCI) guidelines at the primary care level in Burkina Faso. The improvement included a better identification of danger signs and an increase in the proportion of correctly classified children under five years old. A former study reported how FHWs perceived the use and impact of the REC in their daily practice. While a high degree of satisfaction was expressed, FHWs also proposed improvements. FHWs proposed to increase the frequency of supervision and evaluation visits, which usually take place every three months. Supervision from district teams and coaches was globally positively perceived by FHWs, as it allowed them to identify and address errors, and therefore helped them to learn and improve. FHWs also proposed receiving compensations or prizes for the best health centres according to the evaluations.

*Contribution of this study:* In this study, we proposed a method to assess the quality of consultations conducted by FHWs. We focused on anthropometric measurements and vital signs that are systematically measured by FHWs during consultations of children aged between two months and five years old. We showed how this method can feed a live alert system that invites FHWs to verify their input in-real time when potential errors in specific measurements or data entries are identified. We found that height (length) measurements of children were the most challenging, as height (length) was frequently underestimated. Finally, we presented a dynamic dashboard that informs health district managers on the quality of care across the country (using a medal reward system), so they can prioritize their interventions and provide FHWs with targeted support to improve their skills.

## Introduction

The burden of under-five mortality in West Africa is high. Out of 1000 live births in Burkina Faso in 2019, 87 children died before the age of five (1); the leading causes of these deaths are preventable or treatable conditions (2,3). In comparison, the under-five mortality rate in Europe and Northern America was five deaths per 1000 live births in 2019 (4). To reduce the morbidity and mortality of children aged under five years in low- and middle-income countries, WHO and UNICEF developed the Integrated Management of Childhood Illness (IMCI) protocol, which supports the combined treatment of major childhood illnesses (5,6).

The paper-based IMCI was digitalized by Terre des hommes (Tdh) and the Ministry of Health (MoH) in Burkina Faso, who co-created IeDA (the Integrated e-Diagnostic Approach) to improve adherence to IMCI. IeDA includes the Electronic Register of Consultations (REC), a mobile application that runs on Android-based tablets and guides frontline healthcare workers (FHWs) through IMCI to diagnose sick children. The deployment of IeDA in Burkina Faso has been growing steadily, covering 67% of all primary healthcare centres (PHCs) in the country by 2021. It is used by thousands of FHWs during the consultations of millions of children each year.

Using machine learning, important work was done to leverage the large amount of consultation data collected with IeDA (7). IeDA data were analysed to: (i) predict epidemic outbreaks(8,9), (ii) deploy smart dashboards to inform and support decision makers, and (iii) build FHWs capacity to improve the quality of care (10).

During a consultation using the REC, the FHW answers a series of questions related to the child’s anthropometry, vital signs, clinical signs and reported symptoms. The accuracy of the FHW’s answers and his input data are key to a high-quality consultation; the consultation process, the final diagnostic classification and the recommended treatment depend on these data. Very few FHWs who conduct consultations are medical doctors; most of them are nurses, itinerant healthcare workers and midwives. FHWs using the REC are keen to get feedback on their performance and willing to know how they compare to their peers (11). Moreover, managers of PHCs and health districts show a strong interest to be continually informed on how FHWs perform and how they could support their work.

To define what is a high- or low-quality consultation, one needs to define a reference. We can use the REC database itself as a reference and compare each FHW’s input to the typical input over the entire country. This would be equivalent to an anomaly (outlier) detection. However, a weakness of this approach is the possibility of systematic errors which are made by all FHWs collectively and may lead to potential biases. The other possibility, which we use in this study, is to consider an external database consisting of consultations made by experts, which may serve as a reference (12,13).

We focus on anthropometric measurements, which are essential to assess the growth and nutritional status of the child (3,14), and on vital signs (temperature), that are systematically measured by FHWs during consultations of children aged between two months and five years old. We propose a method to quantify the quality of a single consultation and an ensemble of consultations conducted by FHWs. We use this method to feed a live alert system that identifies potential errors in specific measurements or data entries. We explore if this method reveals any bias in the treatment of boys versus girls. Finally, we present a dynamic dashboard that provides managers with an overview on the quality of care across the country so they can support FHW improve their skills.

## Methods

### Data

Data from IeDA includes over nine million consultations, increasing by approximately 200’000 consultations per month. During each consultation, the following information is systematically recorded in real time: age, height (length in case of infants), weight, mid-upper arm circumference (MUAC) and temperature. Z-scores of weight-for-height, height-for-age and weight-for-age are calculated and also recorded. These z-scores are based on the WHO worldwide standard model for growth of children(15). Our main analysis includes data from all consultations conducted during 2020 using the REC, across the country.

To rate the quality of consultations, we use an external database as a reference. We use aggregated statistics from data collected during an earlier project (12,13,16), which we will refer to as `the audit data’ from here on. This audit data was collected between 2014-2017 by specifically trained IMCI experts during the consultations of children aged between two months and five years old, in eight districts (Solenzo, Nouna, Dedougou, Boromo, Toma, Goursi, Ouahigouya and Titao) in Burkina Faso. The aim of this former study was to determine whether IeDA increases adherence to IMCI and hence improves the quality of care for children in PHCs. Due to data-protection, we could not work with the raw audit data. We could however access the means of weight-for-height, height-for-age and weight-for-age z-scores, temperature and MUAC, as well as the covariance matrix of these five variables.

### Reference model

We make the hypothesis that the distribution of patients over the five-dimensional real space of weight-for-height, height-for-age and weight-for-age z-scores, temperature and MUAC is a multivariate Gaussian distribution,

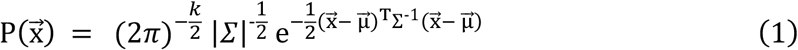

Where 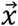 is the *k*-dimensional vector describing a single observation, *k* = 5 is the number of degrees of freedom, Σ is the *k* × *k* positive-definite covariance matrix of the model (estimated from the data), |Σ| is the determinant of the matrix Σ, and 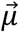 is the vector containing the mean values of the *k* variables. The reference model of ‘good quality of care’ is then fully defined once 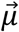 and Σ are known.

### Individual consultations

We first assign a score to individual consultations (data points in the space of anthropometric z-scores, temperature and MUAC); representing the probability that the combination of entered values is accurate. This score allows us to raise a live alert to the FHW when erroneous input data is suspected. With the multivariate Gaussian reference model, the probability density associated with an individual consultation, is given by Equation 1. We trigger an alert when the probability density is lower than a threshold, corresponding to the worst 10% consultations in the database for year 2020. The threshold was set to avoid doing harm to the willingness of FHWs to use the tool at all.

The second step consists in identifying the most suspicious input data in order to guide the FHW in real time to check specific measurements and entries. Therefore, we analyse separately the five variables MUAC, temperature and three z-scores, denoted as *x*_*i*_, with *i* = 1 … 5, and compute individual variable scores score 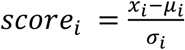, given the reference experts’ means *μ*_*i*_ and standard deviations *σ*_*i*_ Any *x*_*i*_ is suspect when |*score*_*i*_| > *thresh*_*i*_, with *thresh*_*i*_ = 3, ∀ *i* When *score*_*i*_ < -*thresh*_*i*_ or *score*_*j*_ > -*thresh*_*j*,_ *x*_*i*_ is considered extremely low or extremely high, respectively.

When MUAC and temperature are identified as extreme (|*score*_*MUAC*_| > *thresh*_*MUAC*_ and |*score*_*temparature*_| > *thresh*_*temparature*_), the FHW is directly informed by an alert message in the REC and invited to check the input values. Weight, height and age are identified as being abnormally low or high based on the combination of scores related to weight-for-age, height-for-age and weight-for-height (Supplementary Materials).

### Ensembles of consultations over a given time period

#### Probability

We projected consultations 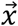 on a new five-dimensional orthogonal space (with uncorrelated dimensions); 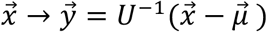, where the column vectors of *U* are the (normalized) eigenvectors of the reference covariance matrix Σ.

For an ensemble of *N* consultations (done by a single FHW, in a single PHC) over a given time period (*N* draws from the reference distribution), the measured sample mean 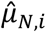 of a single component *y*_*i*_, and its measured sample variance 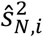 are distributed as Gaussian and 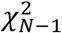 respectively.

The probability density of an ensemble 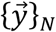 of *N* consultations, is then given by,

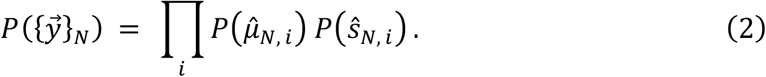

For more details, including equations of 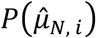 and 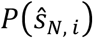, see Supplementary Material. 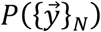 is the probability that the observed ensemble 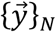 is generated by the reference model. This corresponds to the p-value of the null hypothesis that the consultations are done by an expert. The p-value informs us about the significance of a deviation from the reference model, but it does not necessarily inform us about the quality of the work. In fact, even for the same deviation from the reference model, a large number of consultations will lead to a much lower p-value than a small number of consultations (Figure A 1). Vice versa, different deviations from the reference model can lead to the same p-value, if the number of consultations differs accordingly.

#### Quality score

We aim at providing healthcare workers with an actionable feedback. The feedback consists of two parts: (i) an overall performance score to indicate how the quality of work of a FHW compares to their peers, and (ii) suggestions about which part of their work should be improved with priority. As a proxy for the quality score, we use Equation 2, which defines the probability density that an ensemble of consultations is drawn from the reference distribution (significance), where we fix the number *N* to a constant arbitrary value for all FHWs (Figure A 2). This way, only the mean and variance of the consultation measurements are included in the calculation of the proxy for the quality score; ignoring the number of consultations that gave rise to said mean and variance. To facilitate the interpretation of the score for any user, we define the score as the percentile of a FHW’s score proxy, relative to the entire population of FHWs. The score therefore reflects a percentage-based ranking, and indicates how well the FHW performs compared to their peers.

Figure 1 and Figure A 3 illustrate that the proxy score is more suitable to measure quality, than the actual probability of these ensembles.

**Figure 1.**
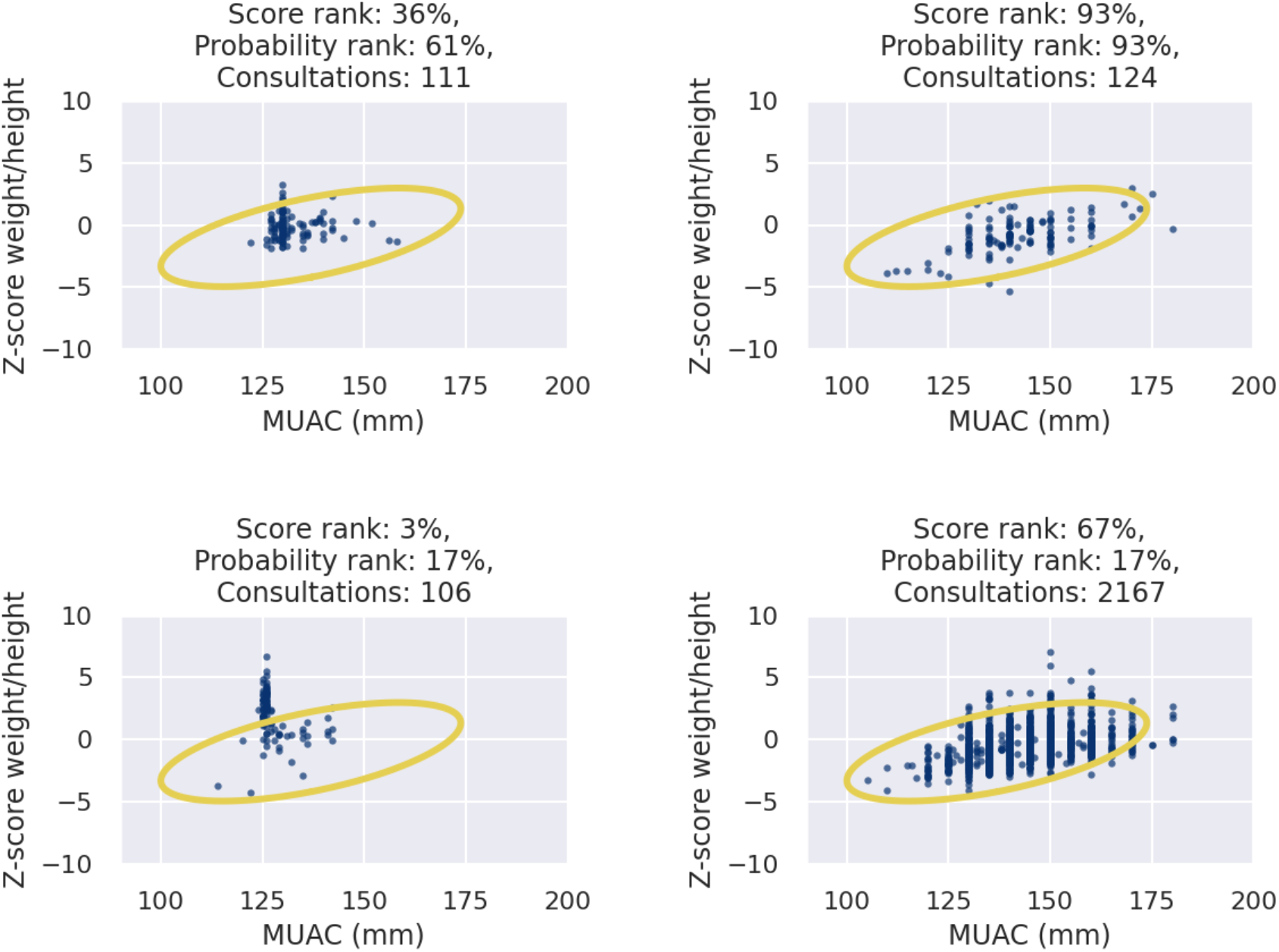
2D projection of the consultations of four Frontline Healthcare Workers (FHWs), over Quarter 4 of 2020. Each dot represents one patient. The yellow reference contour is the 99% confidence level contour of the reference model which was derived from the audit data. Good quality healthcare is reflected by ensembles of consultations which are scattered within the reference contour. In these selected examples, the FHWs on the left clearly make repeated errors in the weight and height measurements of the patients. Our definition of score reflects correctly the difference in quality of care: ranked by score proxy, the FHWs on the left scores the worst (3% on bottom-left; and 36% on top-left), while the FHWs on the right scores best (67% on bottom-right; and 93% on top-right). However, due to the lower number of consultations (n=106) for the FHW in the lower left panel, the probability of its ensemble is the same as for the higher-scoring FHW in the lower right panel (with 2167 consultations): ranked by probability both FHW’s are in the worst 17%. Hence, the true probability is not a useful proxy for the quality score.

### Gender Bias

In order to identify potential gender bias in measurements or data entry, we first compared the typical variation in the scores of individual FHWs over different time periods (quarters of a year), to the typical variation in their scores between boys and girls in the same time period. We also compared the distribution of scores and the frequency of suspicious input values (extremely low/high height, weight, MUAC, etc.) between girls and boys over all FHWs. Statistical significance was assessed using a two-proportion z-test.

## Results

### Detecting common problems

The following analysis includes 2,042,545 consultations (939,761 girls, and 1,202,784 boys) done in 2020, by 7,234 FHWs, in 1,150 PHCs from 39 districts. We compared the distributions of age, temperature, MUAC and the three anthropometric z-scores, i.e., height-for-age, weight-for-height and weight-for-age between the most suspicious consultations, which obtained the lowest scores (worst 0.3%), and the remaining (99.7%) consultations that got higher scores. The distributions are presented in Figure 2. Compared to the consultations with higher scores, the lowest score consultations had lower z-scores of height-for-age (median(IQR) -8.7(−7.5-(−9.8)) vs -2.1(−1.0-(−3.5)), higher z-scores of weight-for-height (median(IQR) 9.0(12.3-6.9) vs -0.2(0.8-(−1.1))), and were more frequent among younger children; 71.6% of consultations with the lowest 0.3% scores involved children under 20 months old vs 40.8% among the remaining 99.7% of consultations. Consultations that involved overweight children (weight-for-age z-score > 2) were also more frequent among the consultations with the lowest scores (15.8% vs 0.5% among consultations with higher scores).

**Figure 2.**
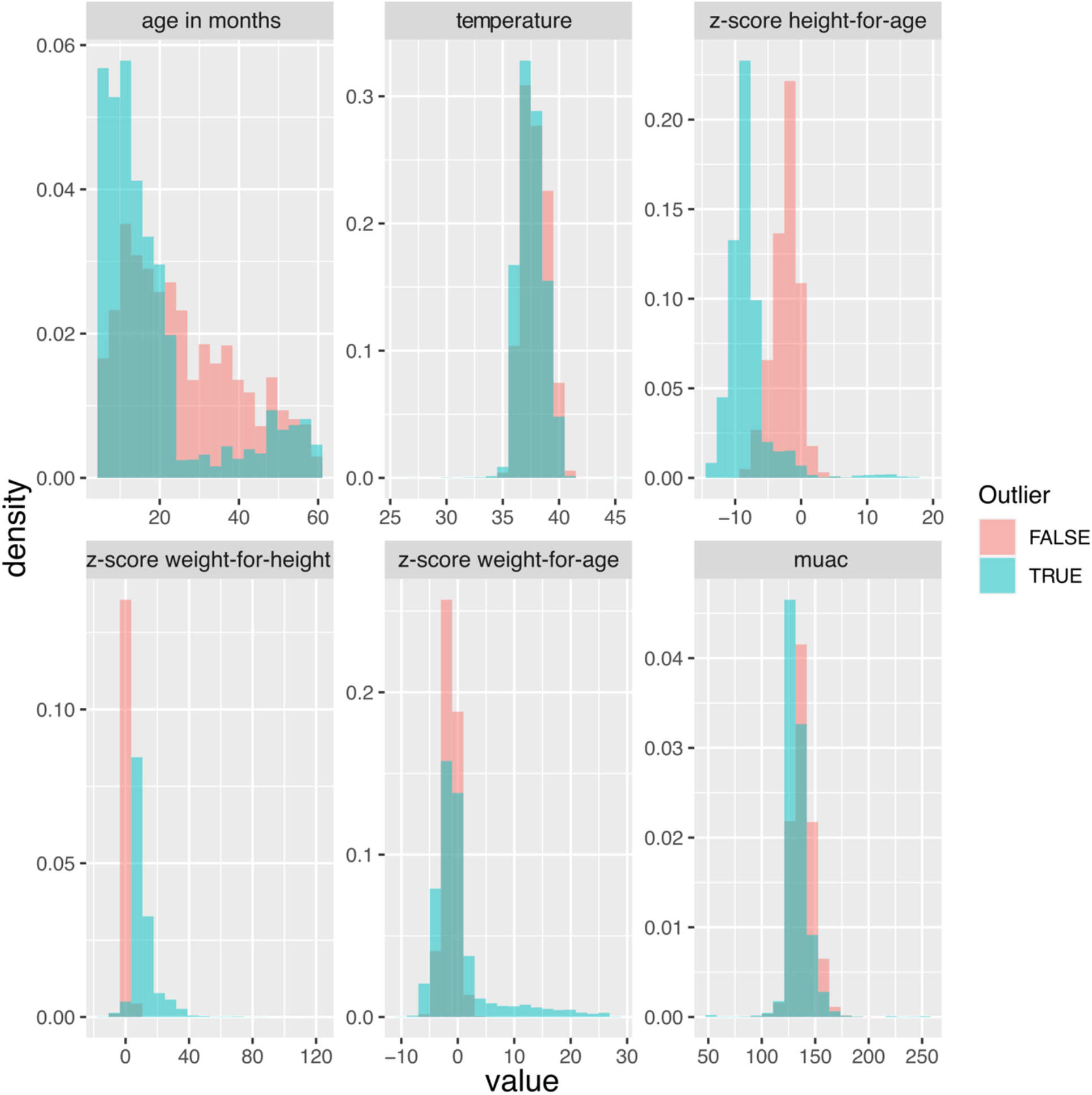
Distribution of age, temperature, MUAC and three anthropometric z-scores among the consultations with the lowest 0.3% scores (denoted as Outlier = TRUE; coloured in blue), and among the remaining 99.7% consultations with higher scores (denoted as Outlier = FALSE; coloured in red).

When identifying the specific inputs (among age, weight, height, MUAC and temperature) with extreme (low or high) values, height underestimation was the most frequent, followed by weight overestimation. Consultations with the 0.3% least likely combinations of anthropometric and vital signs entries underestimated height in 88% of cases and overestimated weight in 14%; see Figure 3 (left). These numbers correspond to 29% of height underestimation and 2% of weight overestimation among the 10% consultations with the lowest scores. Figure 3 (right) presents the evolution of the frequency of underestimated and overestimated input variables as a function of the selected score threshold (varying from the lowest 0.3% to the lowest 40% scores). Reproducing the analysis for consultations conducted in 2019 and 2018 provided similar results (Supplementary Material).

**Figure 3.**
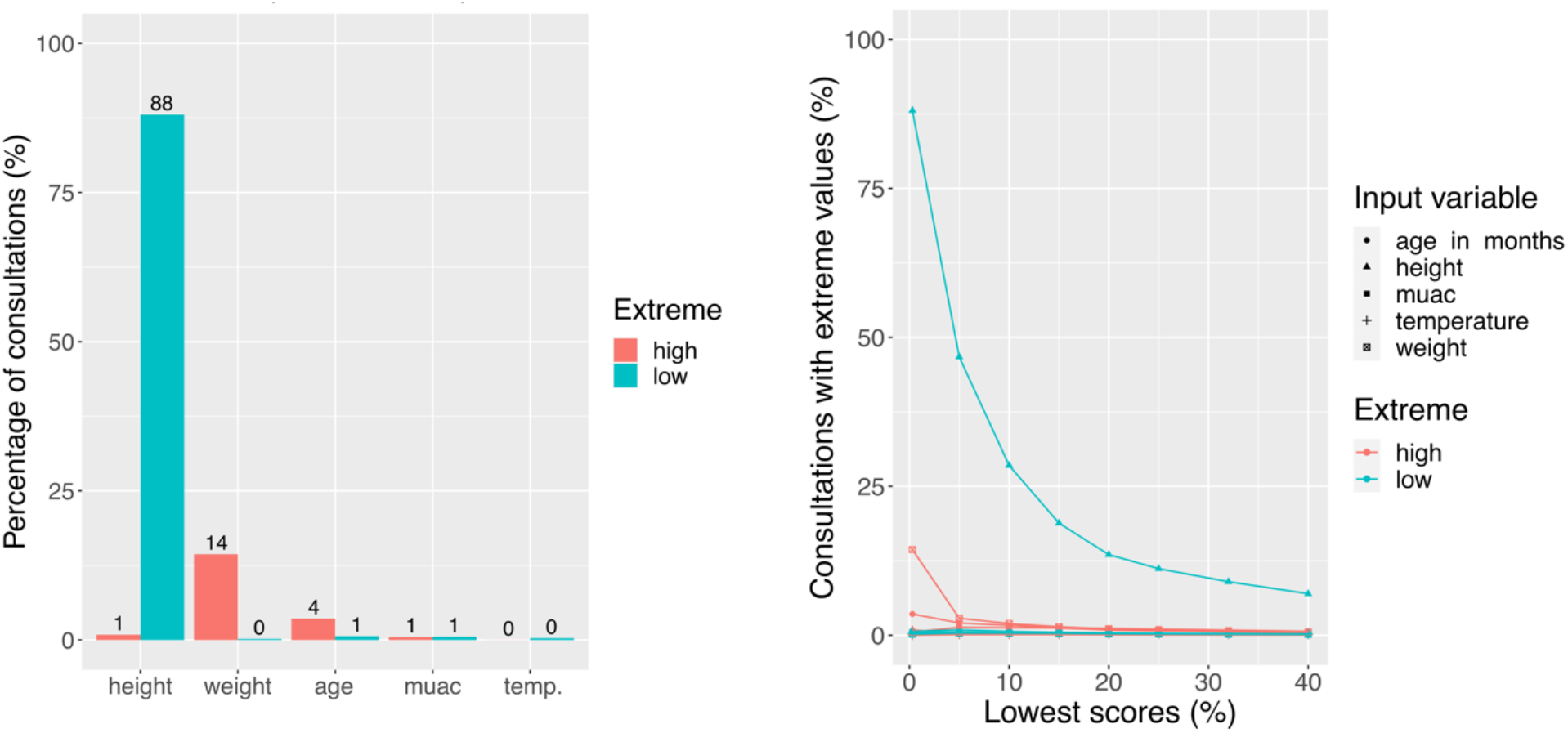
(left) Percentage of extreme anthropometric and vital sign input values among the 0.3% consultations with the lowest scores conducted in 2020 (6,113 /2,042,545 consultations). Blue and red bars represent the percentage of extremely low and extremely high values, respectively. In comparison, we show in (right) the evolution of these percentages among the consultations with the lowest 0.3% to 40% scores.

### Gender bias in primary healthcare

Figure 4 shows that the inter-seasonal variation in quality of care is larger than the variation between boys and girls, as the former is more skewed to the right. We can therefore not demonstrate any systematic gender bias. Considering ensemble of consultation scores of individual FHWs, Figure A 4 shows that only FHWs with fewer consultations had a larger difference in scores between girls and boys. Figure A 5-7 show how the large discrepancy in scores can be explained by outliers having a large impact on the score when the sample size is limited (<1000 consultations).

**Figure 4.**
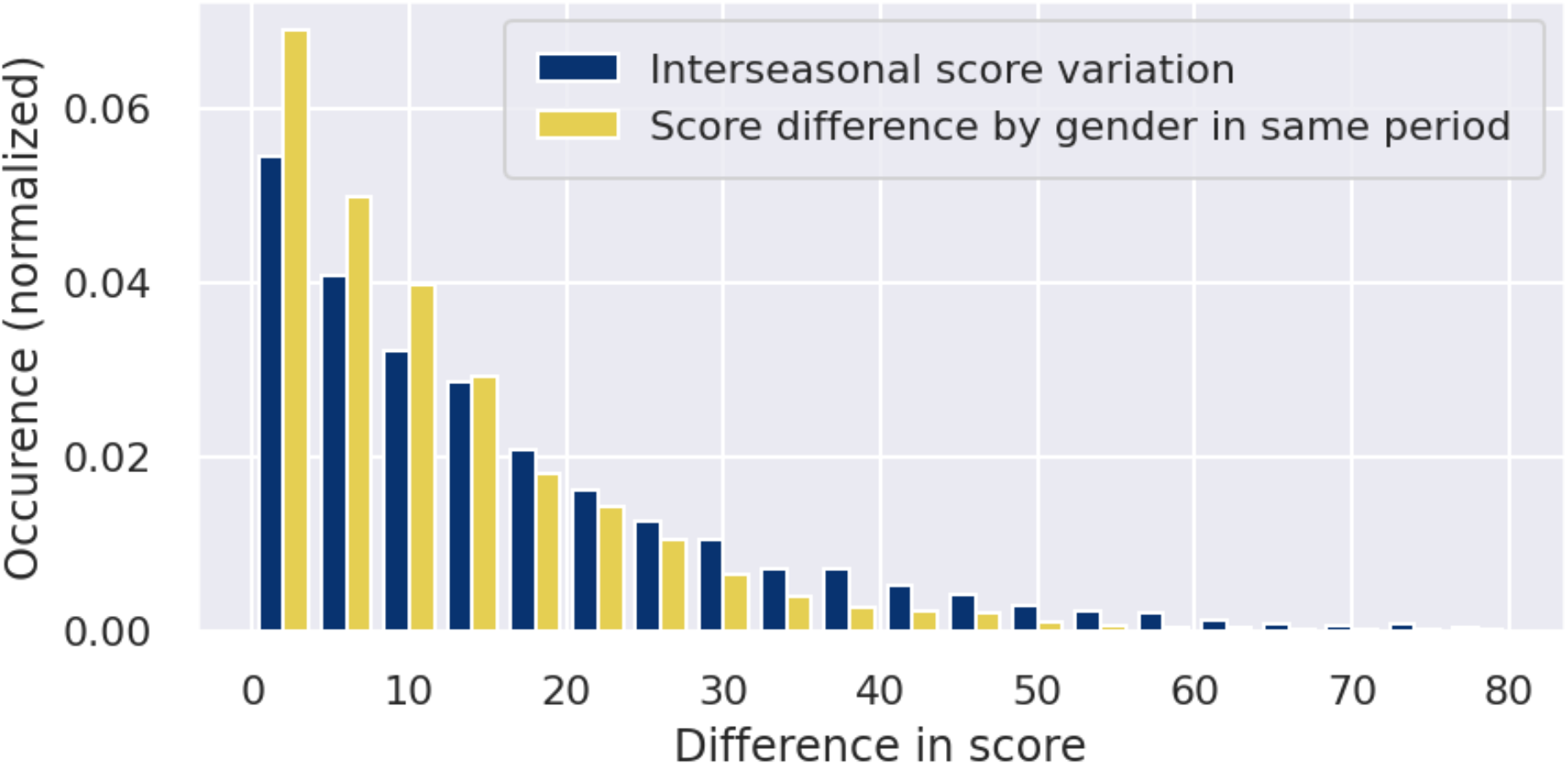
Distribution of absolute difference in scores for the same FHW in different seasons (blue) and for the same FHW in one season but separating the score for boys and girls (yellow). The inter-gender score variation is more skewed toward the right.

Despite the similar overall distribution of scores between girls and boys, the lowest scores for boys were even more extreme (lower) than for girls (see Figure A 8). Among the consultations with the lowest scores (0.3%; 2,818/939,761 girls; 3,316/1,202,784 boys), the most significant difference between the consultations for girls and boys with the lowest scores involved height measurement (Figure 5). Height in girls was less often underestimated than height in boys (84% for girls; 90% boys; p<0.01).

**Figure 5.**
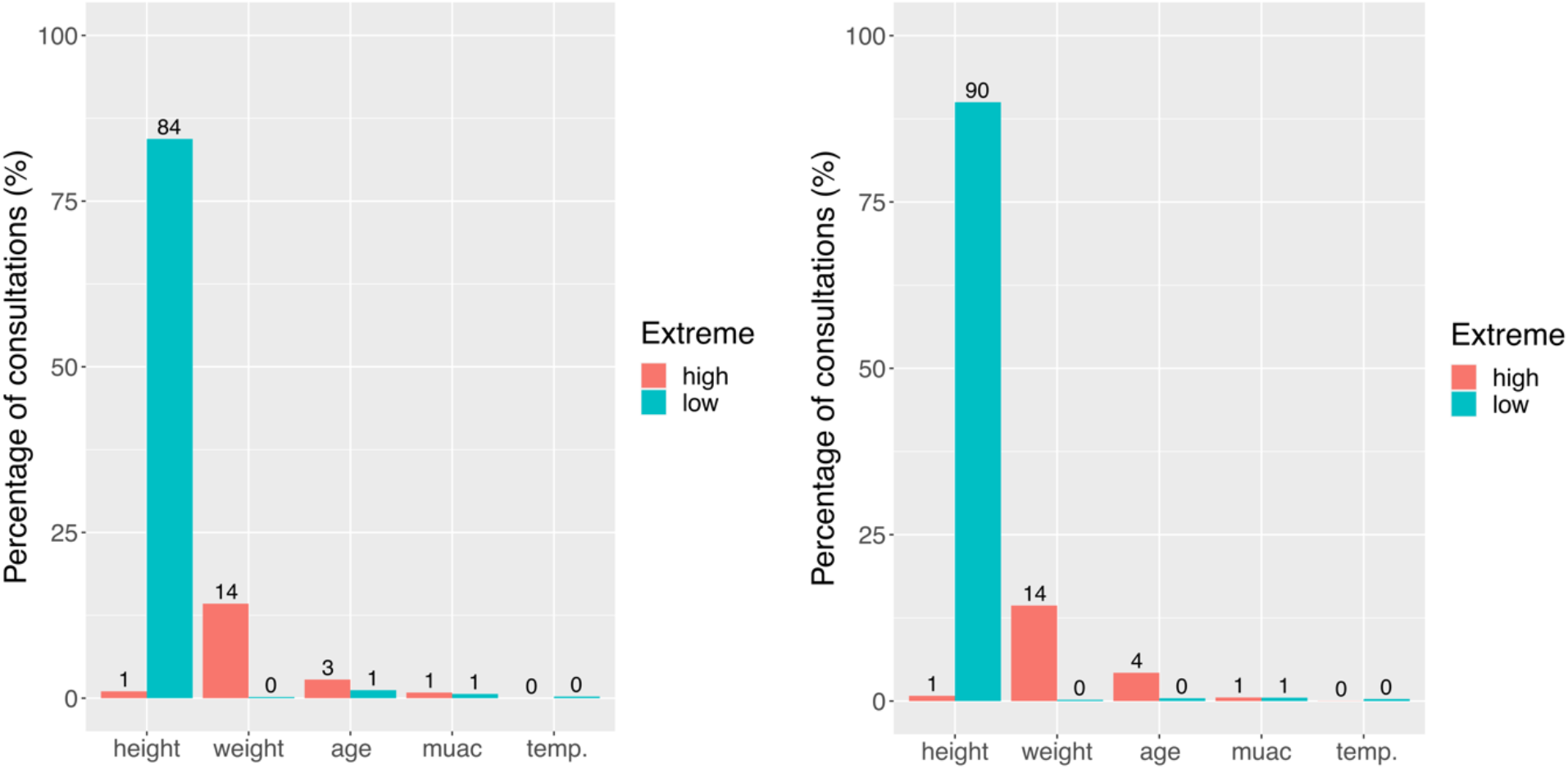
Percentage of extreme anthropometric and vital sign values among (left panel) girls’ consultations (2,818/939,761 consultations), and (right panel) boys’ consultations (3,316/1,202,784 consultations) with the 0.3% lowest scores conducted in 2020. Blue and red bars represent the percentage of extremely low and extremely high values, respectively.

### Live alerts

The REC was expanded to estimate in real time the score of anthropometric and vital signs values that are systematically entered by a FHW during a consultation. When a consultation gets a low score, suspicious values (extremely low or high) are identified, and an alert message invites the FHW to check their entries. In case of a confirmed measurement or data entry error, the FHW can correct the input value and continue the consultation. Figure A 9 presents screenshots of the expanded REC application.

### Dynamic dashboards for managers

A dynamic dashboard was implemented (using Tableau Desktop 2020.3 software) and made accessible online to provide health centre and district managers with an up-to-date overview of the PHC score distributions across the country (see Figure 6). In this dashboard, each PHC was represented by the ensemble of all consultations (focusing on anthropometry and temperature data entries) conducted in this site. Depending on the scores obtained, PHCs were categorized in three categories: gold, silver and bronze (details in Supplementary Material). To help identify the measurements that are the most suspicious in a selected PHC and that lead to a lower quality score, the distribution of all consultations of the PHC are displayed (as a scatter plot) together with the reference model (orange lines) for comparison. In a separate panel, the proportion of gold, silver and bronze medals per health district are also presented. Districts in the Northern and Eastern parts of Burkina Faso (e.g., Dori, Diapaga) had higher percentages of bronze PHCs, while districts in the Western part of the country (e.g., Tougan, Tenado) had a majority of silver and gold PHCs.

**Figure 6.**
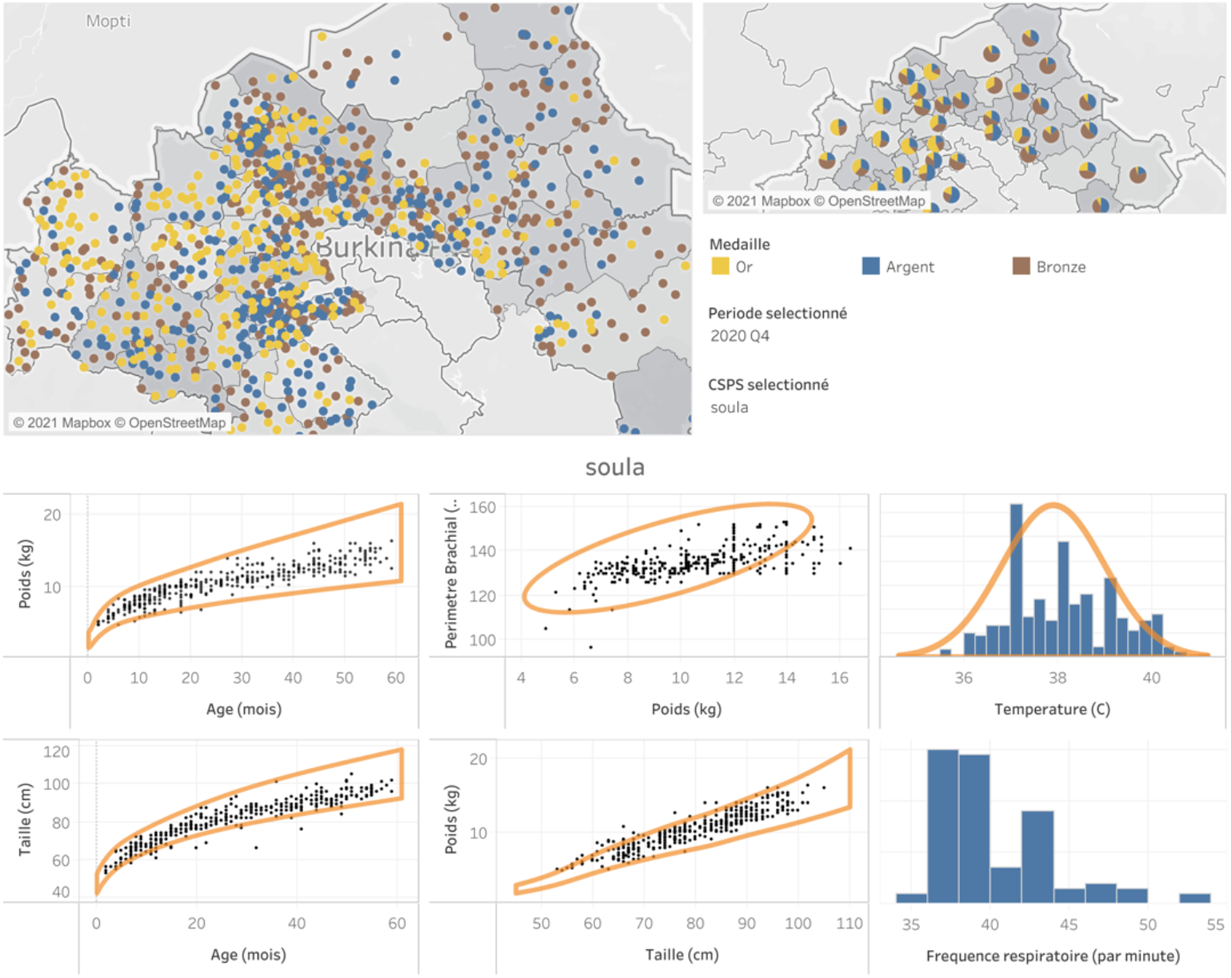
Dynamic dashboard presenting an overview of the PHC scores distributions. Each PHC is represented by the ensemble of consultations that were conducted there during Quarter 4 of year 2020. PHCs are categorized in three categories: gold, silver and bronze based on the obtained score.

## Discussion

Using a multivariate gaussian model, we quantified the quality of anthropometric measures and vital signs as entered by FHWs during consultations of children aged from two months to five years old. The quality measure of individual consultations was used to feed a live alert system that identifies suspect input values of age, height, weight, MUAC and temperature. The quality measure of ensembles of consultations were used to identify suspect behaviours among FHWs based on all the consultations they have conducted during a specific time period.

Among the lowest score consultations, height measurements were the most problematic, as height was frequently underestimated. The challenge when measuring a child’s height (length) lies in the difficulty to keep him fully stretched and still, especially when the child is crying and struggling (19). Among the consultations with the lowest scores, height was significantly more underestimated for boys than for girls, which may be associated to higher activity levels in boys (20–22).

Bronze medals were more frequent in the Northern and Eastern districts of Burkina Faso, while Western Districts had a majority of silver and gold medals. In addition to differences in the quality of anthropometric and vital signs measures, this geographical heterogeneity could also reflect variations in socioeconomic and nutritional status across the country (17,18). Previous reports showed that food insecurity is the most prevalent in the Sahel (Northern) and East regions. If these geographical differences and local child population characteristics are confirmed, the reference model will be adapted accordingly for a more accurate estimation of FHWs’ quality of work.

Our analyses have some limitations. As a reference of what is a high-quality consultation, we used aggregated statistics of experts consultation data (12,13,16). This choice has weaknesses, including that (i) the statistics were computed over a finite number of consultations (order of 1500), and that the reference distribution is only measured up to a few percent accuracy, (ii) the data were collected in a different period than the period in which we tested the quality of care, (iii) the data covered a subset of districts only, (iv) the data were collected by only two trained nurses, which may lead to biased results due to the limited sample size.

In collaboration with the MoH, Tdh has initiated the regular collection of consultations performed by recognized experts, in order to recalibrate the reference model and keep it as close to the real population as possible. Data collection will be done over a heterogeneous sample of the population, and cover different regions and seasons. As we worked with aggregated audit data, we made the simplifying hypothesis that the distribution of patients over the five-dimensional real space of anthropometric and vital signs variables follows a multivariate gaussian distribution. The validity of this assumption and of the audit data will be tested in the future, when the above-mentioned dataset of expert consultations becomes available. As a future work, we will further explore potential factors associated with lower consultation scores (e.g., FHW’s workload, time at which data are entered in the REC), and focus on the quality of reported symptoms and classifications.

## Conclusion

By developing a method that uses a large dataset to qualify the healthcare provided to children in a low-resource setting, we showed how we can foster capacity building through automated personal live feedbacks and dynamic dashboards informing district managers’ decision-making, and explore potential biases in the quality of healthcare.

## Data Availability

The IeDA data may be available with the consent of the Ministry of Health of Burkina Faso.

## Acknowledgements

The authors thank Antoine Geissbuhler, Seydou Toguiyeni, Noël Nacoulma, Noël Zonon, Florian Triclin, Iveth Gonzalez, Sandrine Busiere, Riccardo Lampariello, the IeDA team of Terre des hommes in Burkina Faso and the Ministry of Health from Burkina Faso for their input and fruitful discussions.

