## Supplementary Material for "Improving the quality of anthropometric measures during medical consultations with children aged under five years old in Burkina Faso"

### Real-time identification of extreme input values

Table A2 presents examples of anthropometric and vital signs values reported during single consultations (in 2018), along with their corresponding scores and identified extreme values. These examples were selected among the combinations of entered anthropometric and vital signs values with the 0.3% lowest scores (log-score < -108). A combination of high (low) weight-for-height and low (high) height-for-age suggests a suspicious low (high) input value for height. Similarly, a combination of high (low) weight-for-height and weight-for-age corresponds to a suspicious high (low) input value for weight. Finally, if both weight-for-age and height-for-age are abnormally high (low), this hints to a suspicious low (high) value for age.

| **Age in months** | **Temperature** | **Z-score hfa** | **Z-score wfh** | **Z-score wfa** | **MUAC** | **Log-score** | **Extreme values** |
|---|---|---|---|---|---|---|---|
| 6 | 38,7 | -6,9 | 6,6 | -1,6 | 135 | -184 | Low height |
| 8 | 37,0 | 12,4 | -1,5 | 3,9 | 150 | -116 | Low age |
| 11 | 37,0 | -6,3 | 12,6 | 3,2 | 126 | -209 | Low height; High weight |
| 30 | 38,8 | -0,9 | 19,7 | 12,3 | 140 | -229 | High weight |
| 57 | 39,5 | -6,2 | 8,4 | 0,8 | 190 | -117 | High MUAC; Low height |
| 56 | 41,2 | -8,9 | 8,0 | -1,3 | 141 | -166 | High temperature; Low height |
| 23 | 37,3 | -11,3 | 3,1 | -6,1 | 125 | -157 | High age; Low height |
| 48 | 36,8 | 2,6 | -0,7 | 1,2 | 50 | -120 | Low MUAC |
| 7 | 37,2 | -2,8 | 43,4 | 27,0 | 91 | -1136 | Low MUAC; High weight |
| 23 | 38,0 | 3,4 | 10,4 | 10,3 | 155 | -118 | Low age; High weight |
| 6 | 37,0 | 9,8 | -10,9 | -6,2 | 90 | -412 | Low MUAC; High height; Low weight |

*Table A 1 Examples of anthropometric and vital signs values reported during consultations (in 2018), with their corresponding log-scores and identified extreme values. hfa: height-for-age, wfh: weight for height, wfa: weight-for-age. Variables $x_i$ that are identified as abnormally low with $score_i < -3$ (abnormally high with $score_i > 3$) are highlighted in blue (red).*

### Probability of an ensemble of consultations

For ensembles of consultations, a positive definite matrix can be diagonalized,

$$\Sigma = U \, \Lambda \, U^{-1},$$

$$\Sigma^{-1} = U \, \Lambda^{-1} U^{-1},$$

where the column vectors of $U$ are the (normalized) eigenvectors of the covariance matrix $\Sigma$, and $\Lambda$ is a diagonal matrix with the eigenvalues of $\Sigma$ on the diagonal. As a consequence, the transformation,

$$\vec{x} \rightarrow \vec{y} = U^{-1}(\vec{x} - \vec{\mu}\,),$$

results in uncorrelated Gaussian random variables. By definition $det\ U \equiv 1$, so the Jacobian of the coordinate transformation is 1, such that there is no volume term appearing in the normalization of the probability density $P(\vec{y})$.

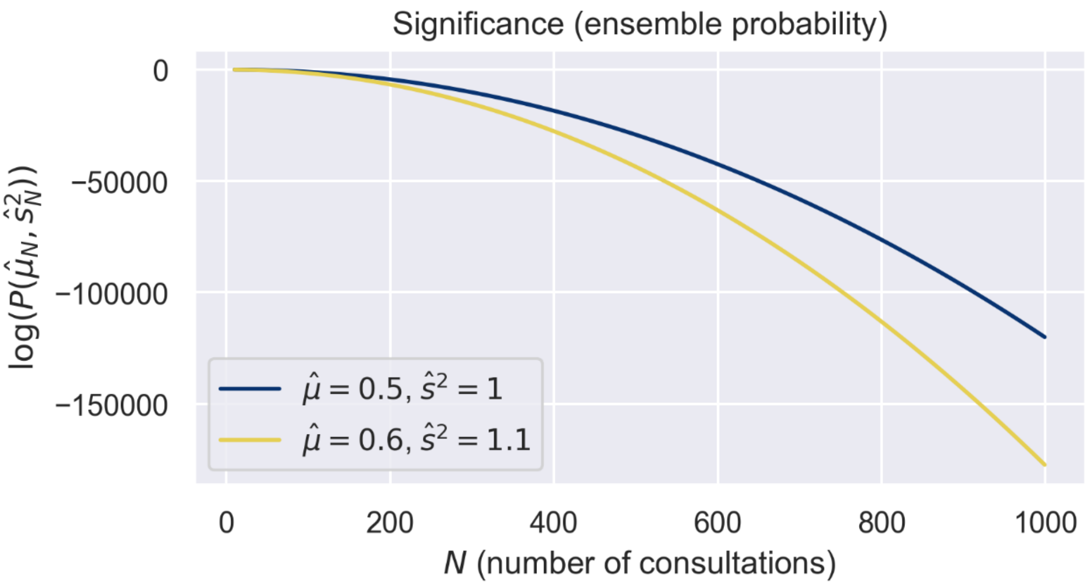


*Figure A 1 Significance (or ensemble probability) as a function of the number of consultations in an ensemble, for two different sample means and sample variances. The range of number of consultations is representative for the range of monthly consultations of children in the different PHC's in the program. The lower (yellow) curve illustrates a slightly larger distance to the reference model. This figure illustrates how the significance is more sensitive to the number of consultations than to the sample mean and variance. For the same mean and variance, a different sample size changes drastically the significance.*

Figure A 1 illustrates the relation between the significance and the size of an ensemble of consultations, and how it is unfit for defining a performance score: all FHWs who recorded over 600 consultations would get a very bad score, regardless of how far they are from the reference.

### Quality of an ensemble of consultations

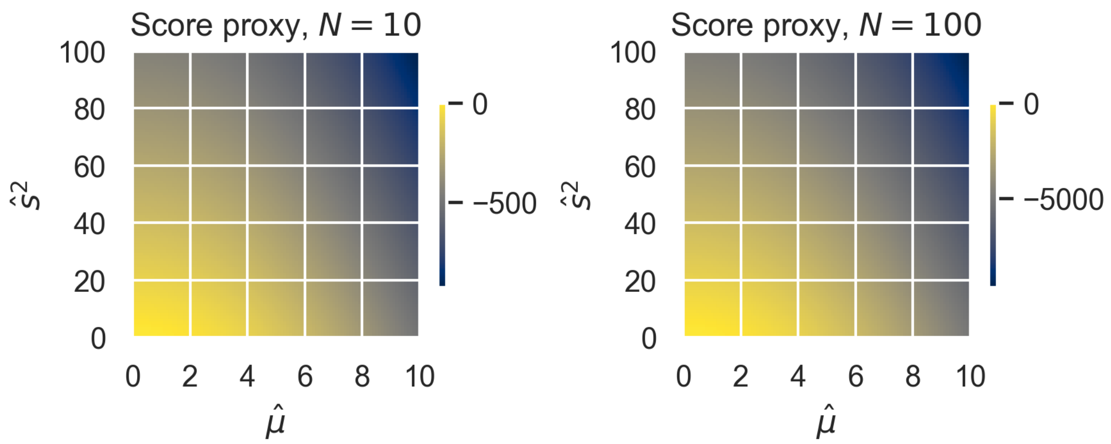


*Figure A 2 Illustration of the score proxy for two different choices of fixed N, ignoring the actual size of an ensemble of consultations. Clearly, since the score is defined as the percentile in which an FHW resides, the choice of N has no impact the final scores, as it has no impact on the ordering of the FHWs.*

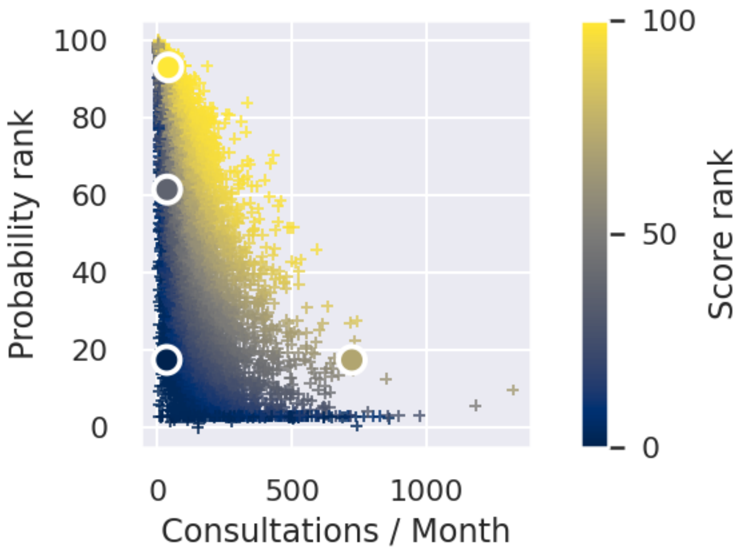


*Figure A 3 The distribution of all FHWs as a function of number of consultations per month, probability of the ensemble of consultations, and the performance score for the same ensemble. A large number of consultations practically excludes a high probability, while allowing for a high score. The four emphasized FHWs (circles), correspond to the four FHWs in Figure 1.*

### Most problematic measurements in 2018 and 2019

Results on the most problematic measurements in 2019 and 2018 were similar to those in 2020. In 2019, among the 0.3% (10%) lowest scores of 1,165,500 consultations, 86% (24%) underestimated height and 18% (3%) overestimated weight. In 2018, for a total number of

1,626,648 consultations, 82% (19%) of the 0.3% (10%) lowest scores underestimated height and 17% (3%) overestimated weight.

### Gender bias

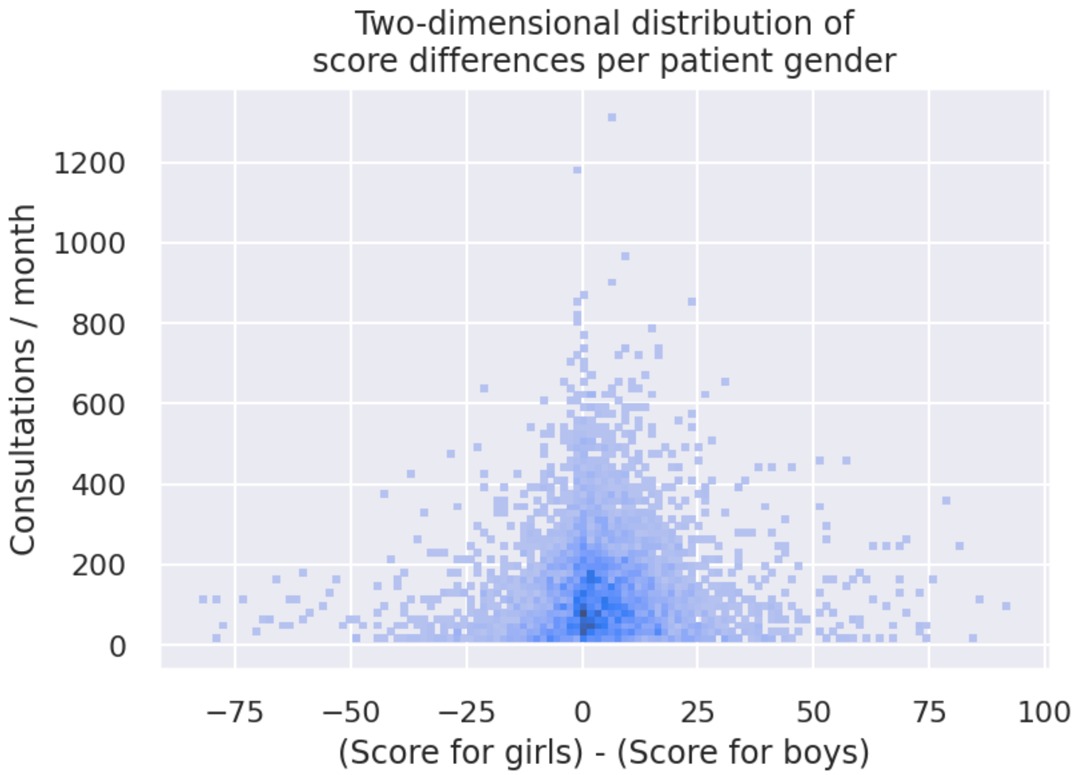


*Figure A 4 Distribution of difference in score when considering consultations of girls or boys only, as a function of total number of consultations. The left of the horizontal axis means a high score for consultations on boys only and a low score for consultations on girls only, and vice versa. Large difference in score only appear in FHWs who generally have fewer consultations to report.*

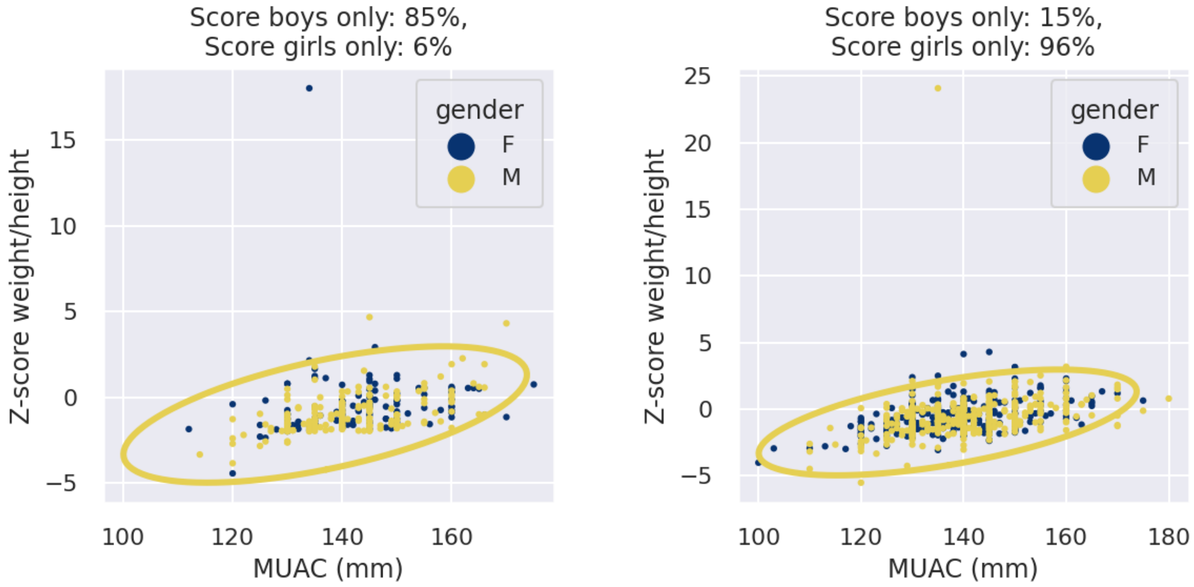


*Figure A 5 Consultations, coloured by gender (yellow for male and blue for female), for two FHWs with a large discrepancy in score when considering only boys or girls. The FHW on the left scores well for boys and bad for girls, and the FHW on the right does the opposite. However, both ensembles are relatively small, and in both cases the difference in score can be entirely attributed to an extreme outlier (in the top of both panels). A single outlier does not justify the conclusion that there is a systematic bias in quality when consulting boys or girls.*

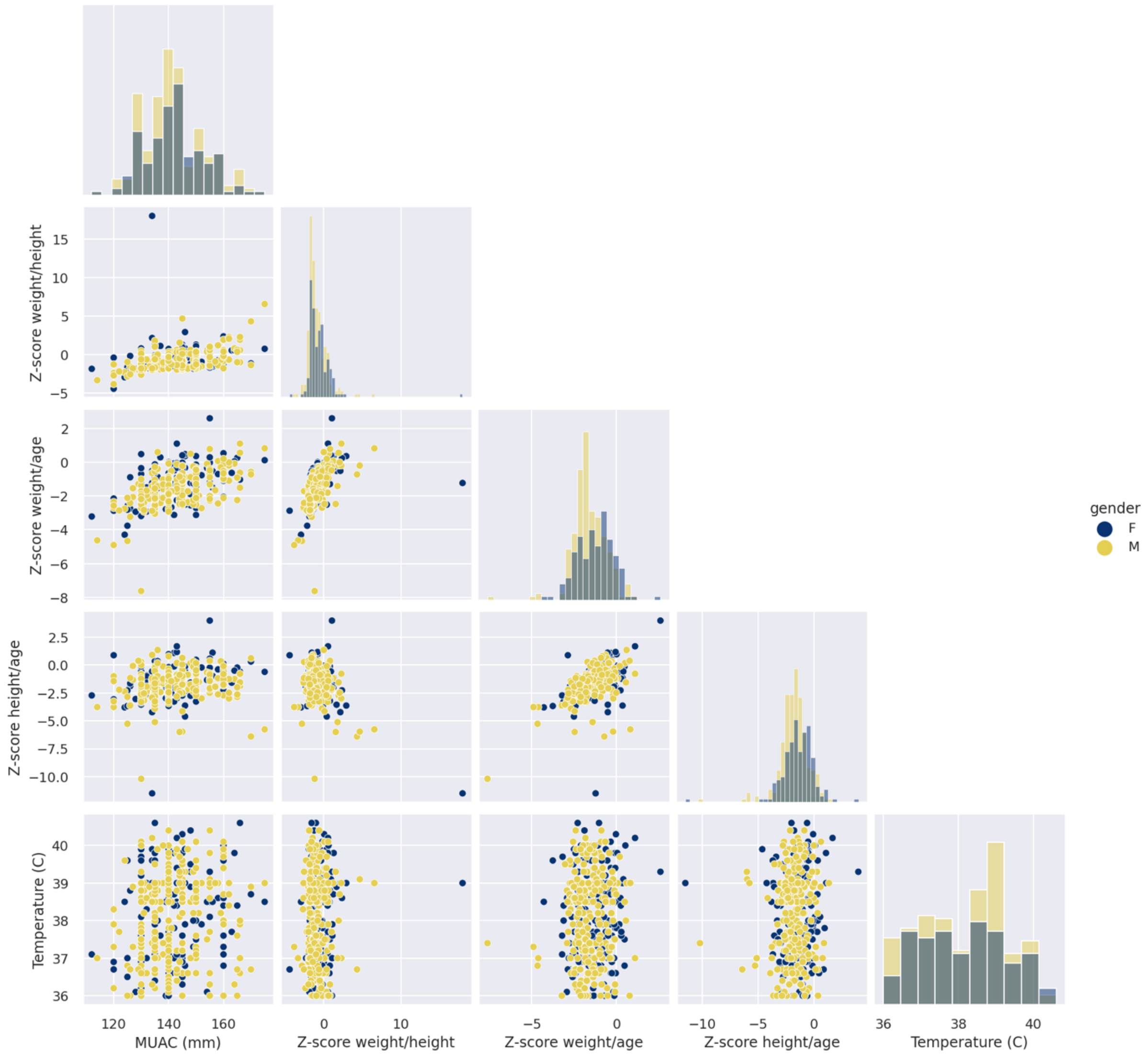


*Figure A 6 The full dimensional representation of the consultations in the left panel of Figure A5, showing that indeed only a handful of outliers is responsible for the big difference in score between boys or girls only.*

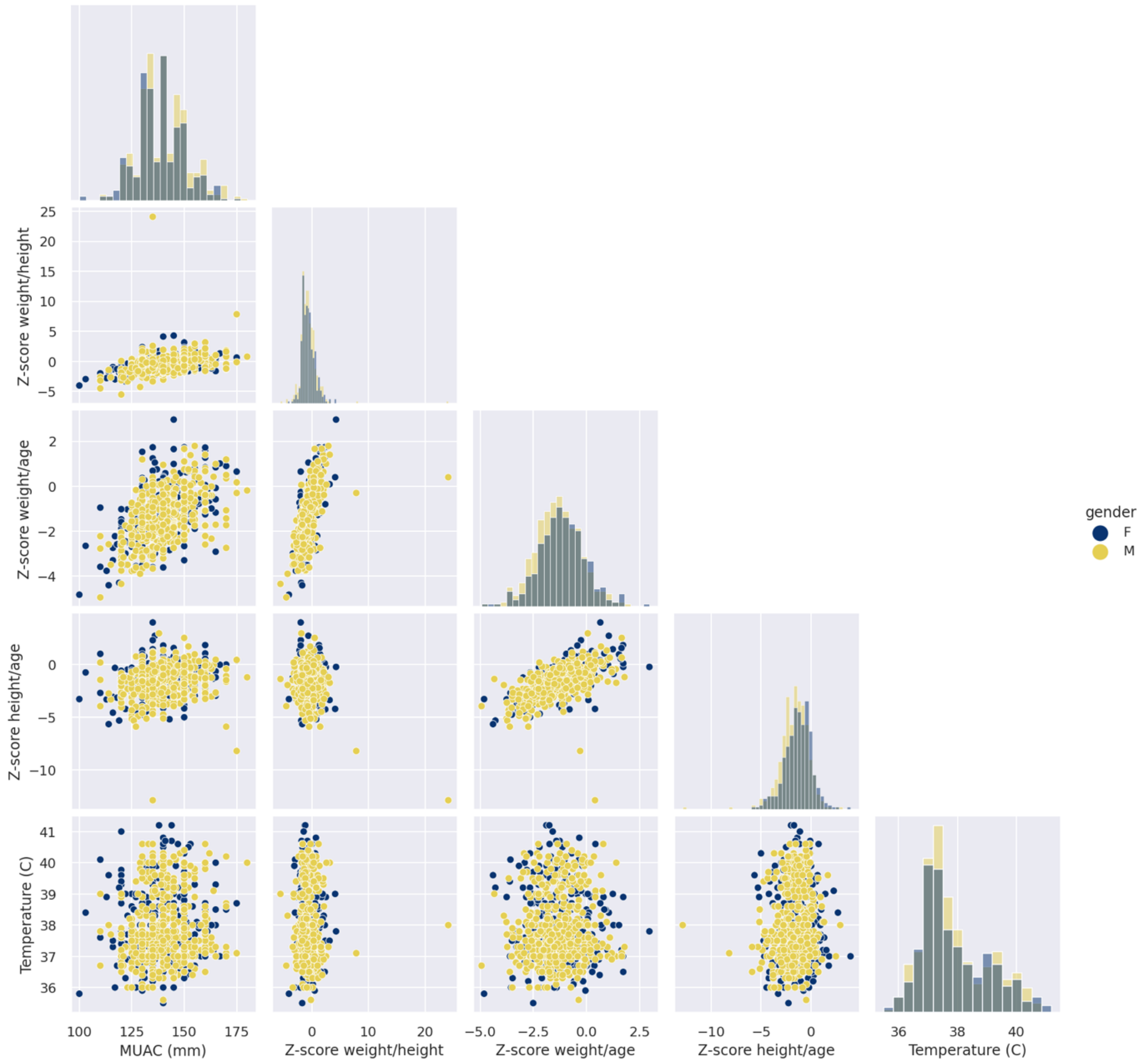


*Figure A 7 The full dimensional representation of the consultations in the right panel of Figure A5, showing that indeed only a handful of outliers is responsible for the big difference in score between boys or girls only.*

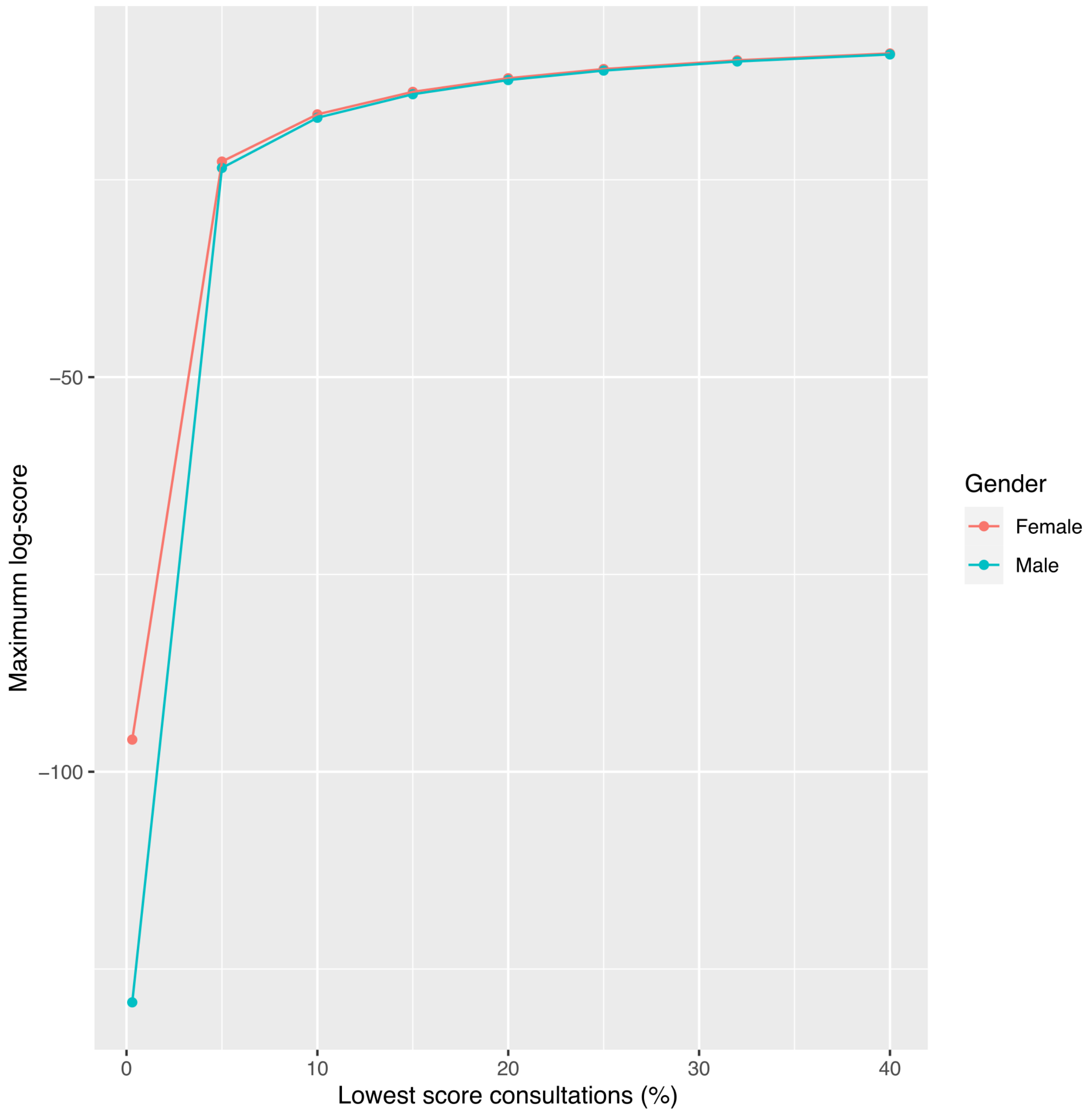


*Figure A 8 Comparison between the scores of girls' (Gender = Female; coloured in red) and boy's (Gender = Male; coloured in blue) consultations. The worst consultations (lowest 0.3% scores) in boys have much lower log-scores than the worst consultations in girls (log-score < -129 for boys vs. -96 for girls).*

### Live alert system and extended REC

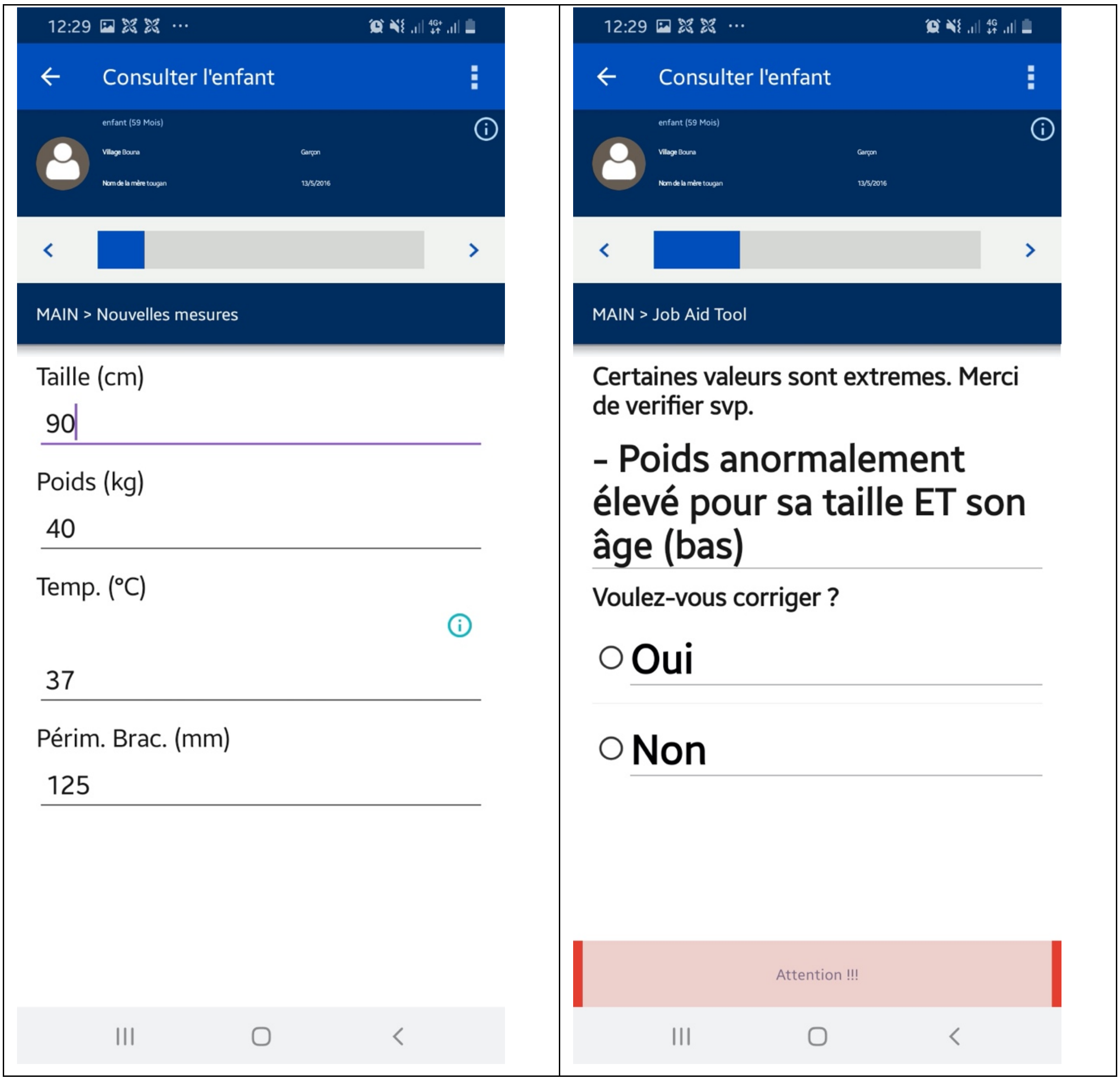


*Figure A 9 Screenshots of the Expanded REC. Based on the input values entered by the FHW (left panel), an alert is raised in real-time and a message invites the FHW to verify specific inputs (right panel). The weight entered is identified to be abnormally high compared to height and age of the child.*

### Dynamic dashboard and PHC medals

The categorization of PHC (gold, silver and bronze) was defined such that in 2020, one third of PHCs, which obtained the best (highest) ensemble of consultation scores, were assigned a gold medal; one third of PHCs, which had the worst (lowest) scores, were assigned a bronze medal; and the remaining PHCs with medium scores were assigned a silver medal. Fixing the score thresholds, which define the three medals and categorise the PHCs accordingly, at a certain point in time (e.g. year 2020) allows us to follow how FHWs’ quality of anthropometric inputs evolve over time
